# Estimation of Effective Reproduction Number for COVID-19 in Bangladesh and its districts

**DOI:** 10.1101/2020.08.04.20168351

**Authors:** Al-Ekram Elahee Hridoy, Mohammad Naim, Edris Alam, Nazim Uddin Emon, Imrul Hasan Tipo, Shekh Md. Shajid Hasan Tusher, Safaet Alam, Mohammad Safiqul Islam

**Affiliations:** Department of Geography and Environmental Studies, University of Chittagong, Chittagong – 4331, Bangladesh; Department of Electrical and Computer Engineering, North South University, Dhaka – 1229, Bangladesh; Integrated Emergency Management and Business Continue Program, Rabdan Academy, Abu Dhabi, UAE; Department of Pharmacy, Faculty of Science and Engineering, International Islamic University Chittagong, Chittagong 4318, Bangladesh; Department of Biochemistry and Molecular Biology, University of Chittagong, Chittagong 4331, Bangladesh; Department of Pharmaceutical Chemistry, Faculty of Pharmacy, University of Dhaka, Dhaka – 1000, Bangladesh; Department of Pharmacy, Faculty of Science, Noakhali Science and Technology University, Noakhali-3814, Bangladesh

**Author notes:** Corresponding author: Mohammad Safiqul Islam, PhD, Professor, Department of Pharmacy, Faculty of Science, Noakhali Science and Technology University, Sonapur, Noakhali-3814, Bangladesh.

**Keywords:** COVID-19, Reproduction number, Effective reproduction number, Bangladesh, Bayesian approach, Time-dependent method

## Abstract

**Background:** Bangladesh is going through an unprecedented crisis since the onset of the COVID-19 pandemic. Throughout the COVID-19 pandemic, the reproduction number of COVID-19 swarmed in the scientific community and public media due to its simplicity in explaining an infectious disease dynamic. This paper aims to estimate the effective reproduction number (Rt) for COVID-19 over time in Bangladesh and its districts using reported cases.

**Methods:** Adapted methods derived from Bettencourt and Ribeiro (2008), which is a sequential Bayesian approach using the compartmental Susceptible-Infectious-Recovered (SIR) model, have been used to estimate Rt.

**Results:** As of July 21, the mean Rt is 1.32(0.98-1.70, 90% HDI), with a median of 1.16(0.99-1.34 90% HDI). The initial Rt of Bangladesh was 3, whereas the Rt on the day of imposing nation-wide lockdown was 1.47, at the end of lockdown phase 1 was 1.06, at the end of lockdown phase 2 was 1.33. Each phase of nation-wide lockdown has contributed to the decline of effective reproduction number (Rt) for Bangladesh by 28.44%, and 26.70%, respectively, implying moderate effectiveness of the epidemic response strategies.

**Interpretation and Conclusion:** The mean Rt fell by 13.55% from May 31 to July 21, 2020, despite easing of lockdown in Bangladesh. The Rt continued to fall below the threshold value one steadily from the beginning of July and sustained around 1. The mean Rt fell by 13.55% from May 31 to July 21, 2020, despite easing of lockdown in Bangladesh. As of July 21, the current estimate of Rt is 1.07(0.92-1.15: 90% HDI), meaning that an infected individual is spreading the virus to an average of one other, with 0.07 added chance of infecting a second individual. This whole research recommends two things- broader testing and careful calibration of measures to keep Rt a long way below the crucial threshold one.

**Highlights:**

- As of July 21, the mean Rt and growth factor is 1.32 and 1.02, respectively.
- Each phase of nation-wide lockdown has contributed to the decline of effective reproduction number (Rt) for Bangladesh by 28.44%, and 26.70%, respectively, implying moderate effectiveness of the epidemic response strategies.
- The Rt of Bangladesh was below 1 for only 20 days, which was observed during May 24- 25, June 19-21, from June 30 to July 6, July 9-12, and July 16-19,2020.
- The initial Rt of Bangladesh was 3, whereas the Rt on the day of imposing nation-wide lockdown was 1.47, at the end of lockdown phase-1 was 1.06, at the end of lockdown phase-2 was 1.33.
- The mean Rt fell by 13.55% from May 31 to July 21, 2020, despite easing of lockdown in Bangladesh.
- The Rt continued to fall below the threshold value one steadily from the beginning of July and sustained around one.
- We suspect that a low testing rate may influence the constant decline of Rt below threshold value 1 in the course of July.
- The mean Rt fell by 13.55% from May 31 to July 21, 2020, despite easing of lockdown in Bangladesh.
- As of July 21, the current estimate of Rt is 1.07(0.92-1.15: 90% HDI), meaning that an infected individual is spreading the virus to an average of one other, with 0.07 added chance of infecting a second individual.

## Introduction

The continued outbreak of COVID-19 in Bangladesh has appeared as a major public health concern and impacted all specters of lives in an unprecedented way. Since its onset in China, various epidemiological projections models have been used globally for the planning and implementation of strategies to mitigate the epidemic. The initial reaction to the COVID-19 pandemic in Bangladesh was closing down schools, universities and locking down the whole country as a measure for preventing the virus from spreading. Since COVID-19 is evolving in an unprecedented nature, researchers have been using various models to predict the final size of the number of cases, dynamics, and peak-time. In the Bangladesh context, Hridoy et al.^1^ used the Logistic Growth model to predict the number of cases, peak-time, and a plausible endpoint of the COVID-19 epidemic in Bangladesh. However, the number of new cases over time referred to as incidence, acts as a proxy for risk but does not act as a metric for the epidemic progression or prediction of its evolution.^2,3^ Among the various metrics that depend on daily changes data, one metric is most promising and could capture potential disease progression. The metric is known as, *R*_0_ which denotes the number of secondary infections generated by a single infection and is often used as a metric to measure transmissibility of an infectious disease.^3^ An *R*_0_ value 3 represents that on average, each infected person has been spreading the infection to 3 others. The *R*_0_ can be estimated at the beginning of an epidemic and its constant when no immunity intervention measures are implemented, When an epidemic progresses a more practical and an effective version of *R*_0_, the Rt can be estimated, which refers to the number of susceptible people in a population who can be infected by an infected individual at any given time. Although the basic reproduction number *R*_0_ is useful in understanding the transmissibility of a disease; it assumes that the epidemic first occurs in a fully susceptible population; hence, it is important to evaluate time-dependent variations in the transmission potential of infectious diseases.^4^ An effective reproduction number (Rt) captures the impact of preventive measures such as quarantine and isolation protocols, travel restrictions, school closures, physical distancing, the use of face masks that are put in place to contain an infectious virus. Since the Rt values change place to place, understanding the gravity of it at a nation and sub-national level, decision-makers can more precisely adjust their intervention strategies. Besides, it is an important parameter to evaluate the effectiveness of current public health interventions and plans additional interventions as required.^5^ When Rt =1, the epidemic is steady, If Rt > 1, the number of cases will increase exponentially and Rt < 1 means the epidemic will slow down and likely to fade out since an infected person less infect another person. The Rt number would typically be less than the *R*_0_ number as the effect of control measures and non-susceptible people in the population are taken into account.^6-8^ The effective reproductive number (Rt) is a useful metric that can be used to follow-up the epidemics since it can evaluate the potential impact of the intervention in the course of a pandemic.^6,9,10.^ The time-varying real-time effective reproduction number (Rt) gives policy-makers an up-to-date snapshot of the current epidemic situations and helps to know, when, and where stringent policies are needed. The epidemic situation becomes manageable when the Rt below stays a crucial threshold below 1 long way.

Thus, understanding and managing the time-varying Rt of Bangladesh and its district is of great importance for policy-makers; otherwise, Bangladesh will be flying blind in the course of this epidemic. The key parameters to measure the transmissibility of an infectious disease can be estimated through the distribution of the generation interval time which refers to the time difference between infection events in an infector-infectee pair, serial interval (time between symptom onsets in an infector-infectee pair) and incubation period (time between the moment of infection and symptom onset).^11,12^ Zu et al.^13^ estimated the mean serial Interval for Wuhan was 7.5 days (95% CI: 5.3–19). Nishiura et al.^14^ estimated the median serial interval (SI) for COVID-19 is 4.0 days (95% CI: 3.1, 4.9) and also found that SI of COVID-19 is shorter than its median incubation period which suggests that a substantial proportion of secondary transmission may occur prior to illness onset. A recent analysis by^15^ suggests the mean interval was 3.96 days (95% CI 3.53-4.39 days), where the authors used the 468 COVID-19 samples that reported in mainland China as of early February. Inglesby^16^ determined the Rt as an indicator to measure the transmission of SARS-CoV-2 before and after the interventions and they estimated in early through mid-January 2020, the SARS-CoV-2 epidemic in Wuhan had an Rt of 3 to 4. Zu et al. ^13^ estimated in late January the effective reproduction number was 2.620 (95% CI: 2.567–2.676) and had dropped below 1.0 since February 5, 2020. You et al. ^17^ estimated effective reproduction rate for Hubei Province and outside of Hubei provinces, for Hubei province, Rt had dropped below 1 around February 19, 2020, and outside Hubei Province, Rt dropped below 1 around February 11, 2020, which indicated that the containment measure carried out by Chinese government was effective and efficient.

The first three COVID-19 cases were reported in Bangladesh on 8^th^ March 2020. It spread to the maximum of the districts of the country within a month. After the confirmation of the first three cases of COVID-19 on 8^th^ March 2020, the government of Bangladesh declared a ten days lockdown from 26^th^ March 2020 to 4th April 2020. However, lockdown periods were later extended several times until 30^th^ May 2020, and then zone-based lockdown strategies have been taken. Since there is no country-wide lockdown in place now, the potential impact of COVID-19 in Bangladesh is still unknown. Therefore, given the immediacy of the COVID-19 situation, this paper aims to estimate the effective reproduction number (Rt) of COVID-19 for an ongoing epidemic in Bangladesh on a daily and weekly basis and its districts using reported data (diagnostic testing/ reported cases). Understanding the recent Rt for COVID-19 of Bangladesh and its district will help policy-makers to take stringent policies to slow the spread of virus and understanding Rt for COVID-19 of Bangladesh and its districts could reveal how effective and efficient the undertaken stringent policies were during the epidemic. The study highlights the importance of the discovery of disease transmission and the necessity of mitigation measures to reduce COVID-19 transmission in Bangladesh.

## Method

### Dataset

The COVID-19 data used in this research is collected from the Institute of Epidemiology Disease Control and Research (IEDCR) and Directorate General of Health Services (DGHS). We have used district-wise COVID-19 data from 15^th^ April 2020 to 4^th^ July 2020, and for overall Bangladesh, we have used data from 14^th^ March 2020 to 20^th^ July 2020, since there were no new cases identified during 8^th^ March 2020 (1^st^ reported case) - 14^th^ March 2020. Analyses for few districts were absent due to incomplete or inconsistent and low number of reported cases from 15^th^ April. The calculations and visualizations were generated using NumPy, pandas, matplotlib, SciPy stats and interpolation packages of Python 3.

### Serial Interval

As mentioned earlier, the transmissibility of an infectious disease can be estimated through the distribution of the generation interval time which refers to the time difference between infection events in an infector-infectee pair, serial interval (time between symptom onsets in an infector-infectee pair) and incubation period (time between the moment of infection and symptom onset) ^11,12^. According to a recent analysis by Du et al. ^15^, the mean interval was 3.96 days (95% CI 3.53-4.39 days), where the authors used the 468 COVID-19 samples reported in mainland China as of early February. Nishiura et al. ^14^ estimated the median serial interval(SI) for COVID-19 is 4.0 days (95% CI:3.1, 4.9) and also found that SI of COVID-19 is shorter than its median incubation period which suggests that a substantial proportion of secondary transmission may occur prior to illness onset and Significant pre-symptomatic transmission would probably reduce the effectiveness of control measures that are initiated by symptom onset.^18^ Wei et al. ^19^ reported that an investigation of all 243 cases of COVID-19 reported in Singapore between January 23 and March 16, 2020, and identified seven clusters of cases where pre-symptomatic transmission was the most probable cause of secondary cases. Moreover, asymptomatic carriers can transmit SARS-CoV-2. ^20^ IEDCR found that 23% of all Covid-19 cases were asymptomatic in Bangladesh. ^21^

### Calculation of real-time *R*_*t*_

We can calculate real-time *R*_*t*_ by using the Bayesian approach proposed by Bettencourt & Ribeiro ^22^ and adapted by data machine learning technology.^23^ We have access to the data of COVID-19 new cases on a daily basis which gives us a clue about the current value of *R*_*t*_. Moreover, today’s value of *R*_*t*_ is related to the value of yesterday’s value *R*_*t*−1_ and thus every previous value of *R*_*t*−*m*_ for that matter. Bayes’s rule updates the beliefs about the true value of *R*_*t*_ based on the information of how many new cases have been reported each day.

Bayes’ Theorem suggests,

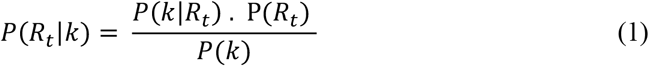

Here,

*P*(*k*|*R*_*t*_) is the likelihood of observing *k* new cases given *R*_*t*_ time,

P(*R*_*t*_) is the prior beliefs of the value P(*R*_*t*_) at the beginning of the study period,

*P*(*k*) is the probability of observing *k* new cases for a given day.

This calculation is for a single day. To calculate everyday data, we use yesterday’s prior P(*R*_*t*−1_) to estimate today’s prior P(*R*_*t*_). We will assume an uncorrelated Gaussian process, so P(*R*_*t*_|*R*_*t*−1_) = 𝒩(*R*_*t*−1_, σ), where s is a hyperparameter.

So, on day one:

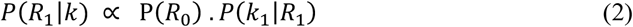

On day two:

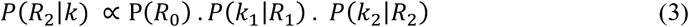

And so on.

### Choosing a Likelihood Function *P*(*K*_*t*_|*R*_*t*_)

A likelihood function says how likely we are to see *k* new cases, given a value of *R*_*t*_. Given an average arrival rate of 𝜆 new cases per day, the probability of observing *k* new cases is distributed according to the Poisson Distribution:

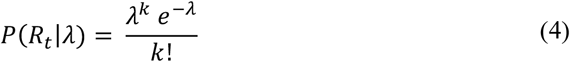

And the relationship between *R*_*t*_ and 𝜆:

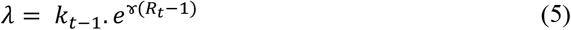

Where ɤ is the reciprocal of the serial interval and according to recent findings by Du et al. ^15^, the mean serial interval of four days has been considered for this research.

Further, new cases are known; therefore, the likelihood function as a Poisson parameterized by fixing*k* and varying *R*_*t*_ can be reformulated.^22^

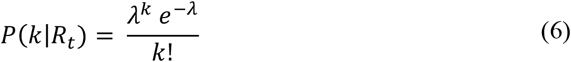

Moreover, to address erratic case reporting due to testing backlogs, a gaussian filter has been applied to the time series to get true data.

## Highest Density Interval (HDI)

The HDI indicates which points of distribution are most credible, and which cover most of the distribution. Thus, the HDI summarizes the distribution by specifying an interval that spans most of the distribution, of 95% of it, every point inside the interval has higher credibility than any point outside the interval. ^24^ Some authors proposed that 95% may not be the most appropriate for posterior Bayesian distributions, potentially lacking instability if not enough posterior samples are taken. ^24^ The proposition was to use 90% rather than 95%. Thus, in this research, 90% HDI has been considered.

### Growth Factor

Growth Factor is another metric to capture how cases change over time. The growth factor is the variable by which a quantity multiplies itself over time. When a growth factor > 1, it indicates an increase of cases, while it is remaining between 0 and 1, it is a sign of decline, when a growth factor persistently > 1 could signify exponential growth.

The growth factor for a certain day is defined as below:

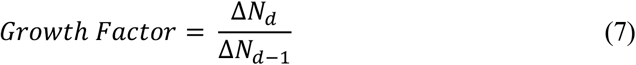

Where,

*N*_*d*_ is the active cases count on the current day,

*N*_*d*−1_ is the active cases count on the previous day.

## Result

According to the number of diagnosed cases from 8th March to 15 July, the real-time effective reproduction number (Rt) has been estimated. The government of Bangladesh started to impose country-wide lockdown from 26^th^ March 2020, before imposing nation-wide lockdown the average effective reproduction number (*R*_*t*_) over seven days was 2.58 (1.06-4.68: 90% HDI) from March 13- 20, 2020, which is presented in Table 1. After imposing lockdown, the *R*_*t*_ plummeted to 1.41 (0.44- 2.37: 90% HDI) in the 3^rd^ week, but scenarios got worsen in the 4^th^ week, the mean weekly *R*_*t*_ increased to 2.20 (1.62-2.82: 90% HDI), which is a 56% increase from the 3rd week. It is observed that in the 16^th^ week, 17^th^ week, and 18^th^ week the mean weekly *R*_*t*_ dropped below the *R*_*t*_ threshold 1. It is also observed that weekly mean *R*_*t*_was started to decline from 13^th^ week to 17^th^ week consecutively and again ramped up in 18^th^ week.

**Table 1:** Time-varying effective reproduction number for Bangladesh.

| **Week** | **Date** |  | **ML** | **Low_90** | **High_90** | **Relative Changes (%)** |
| --- | --- | --- | --- | --- | --- | --- |
|  | **From** | **To** |  |  |  |  |
| **1** | 2020-03-14 | 2020-03-20 | 2.58 | 1.06 | 4.68 |  |
| **2** | 2020-03-21 | 2020-03-27 | 1.66 | 0.68 | 2.71 | -35.65 |
| **3** | 2020-03-28 | 2020-04-03 | 1.41 | 0.44 | 2.37 | -15.06 |
| **4** | 2020-04-04 | 2020-04-10 | 2.20 | 1.62 | 2.82 | +56.02 |
| **5** | 2020-04-11 | 2020-04-17 | 1.74 | 1.43 | 2.06 | -20.9 |
| **6** | 2020-04-18 | 2020-04-24 | 1.25 | 1.02 | 1.50 | -28.16 |
| **7** | 2020-04-25 | 2020-05-01 | 1.18 | 0.97 | 1.41 | -5.60 |
| **8** | 2020-05-02 | 2020-05-08 | 1.14 | 0.95 | 1.34 | -3.38 |
| **9** | 2020-05-09 | 2020-05-15 | 1.22 | 1.05 | 1.40 | +7.01 |
| **10** | 2020-05-16 | 2020-05-22 | 1.23 | 1.08 | 1.39 | +0.81 |
| **11** | 2020-05-23 | 2020-05-29 | 1.10 | 0.96 | 1.24 | -10.56 |
| **12** | 2020-05-30 | 2020-06-05 | 1.16 | 1.04 | 1.28 | +5.45 |
| **13** | 2020-06-06 | 2020-06-12 | 1.09 | 0.98 | 1.20 | -6.03 |
| **14** | 2020-06-13 | 2020-06-19 | 1.06 | 0.96 | 1.17 | -2.75 |
| **15** | 2020-06-20 | 2020-06-26 | 1.02 | 0.92 | 1.13 | -3.77 |
| **16** | 2020-06-27 | 2020-07-03 | 0.95 | 0.85 | 1.05 | -6.86 |
| **17** | 2020-07-04 | 2020-07-10 | 0.93 | 0.82 | 1.04 | -2.10 |
| **18** | 2020-07-11 | 2020-07-17 | 0.97 | 0.86 | 1.09 | +4.30 |

Based on Figure 1, It is observed that Bangladesh attained a maximum value of *R*_*t*_ 2.44 (1.78, 3.1: 90% HDI) on 6^th^ April 2020 (Table 2). The red dots represent the most likely value of Rt, and the shaded grey area is the 90% Highest Density Interval (HDI). During the initial stage of the epidemic, Rt fluctuated within the range of 3.00-1.61(from March 14^th^ to 25^th^ March 2020). After the first intervention phase, Rt fluctuated within the range of 2.44-1.02(from 26^th^ March to 25 April 2020). According to Figure.1, a significant increase of Rt has been observed during April 3-15, 2020. After the second extended intervention phase (from 26^th^ April to 30^th^ May), Rt fluctuated within the range of 1.33-1.06. The government of Bangladesh lifted the nation-wide lockdown on 31^st^ May 2020, in this period, despite easing of lockdown, Rt fluctuated within the range of 1.27-0.82 and mean Rt was 1.02. It is observed that for the first time on May 24-25,2020, Rt was below 1, which was around four months later since the outbreak. It is observed that during June 19-21, 2020 for 3 consecutive days and from June 30 to July 6, 2020, for seven consecutive days, Rt dropped below threshold value 1 and again dropped below 1 during July 9-12 and July 16-19 for 4 consecutive days and reached above 1 on 20^th^ July 2020. The mean Rt for COVID-19 in Bangladesh from March 14, 2020, to July 21, 2020, is 1.32(0.98-1.70, 90% HDI), with a median of 1.16(0.99-1.34 90% HDI), which is presented in Table 2. According to Figure.1, at the initial phase of the outbreak (on March 14, 2020), the Rt was 3 (1.01-6.99: 90% HDI).

**Table 2:** Descriptive statistics for effective reproduction number (Rt)

| *Lockdown Status* | *Time-period* |  | *Mean* | *Median* | *Maximum* | *Minimum* | *IQR* | *Relative Change (%)* |
| --- | --- | --- | --- | --- | --- | --- | --- | --- |
|  | *From* | *To* |  |  |  |  |  |  |
| *No-lockdown* | 2020-03-14 | 2020-03-25 | 2.25 | 2.13 | 3.22 | 1.61 | 0.82 | - |
| *1st* | 2020-03-26 | 2020-04-25 | 1.61 | 1.51 | 2.44 | 1.02 | 0.55 | -28.44 |
| *2nd* | 2020-04-26 | 2020-05-30 | 1.18 | 1.19 | 1.33 | 0.95 | 0.12 | -26.70 |
| *No-lockdown* | 2020-05-31 | 2020-07-21 | 1.02 | 1.03 | 1.27 | 0.82 | 0.13 | -13.55% |
| *Overall* | 2020-03-14 | 2020-07-21 | 1.32 | 1.16 | 3.22 | 0.82 | 0.31 |  |

**Figure 1:**
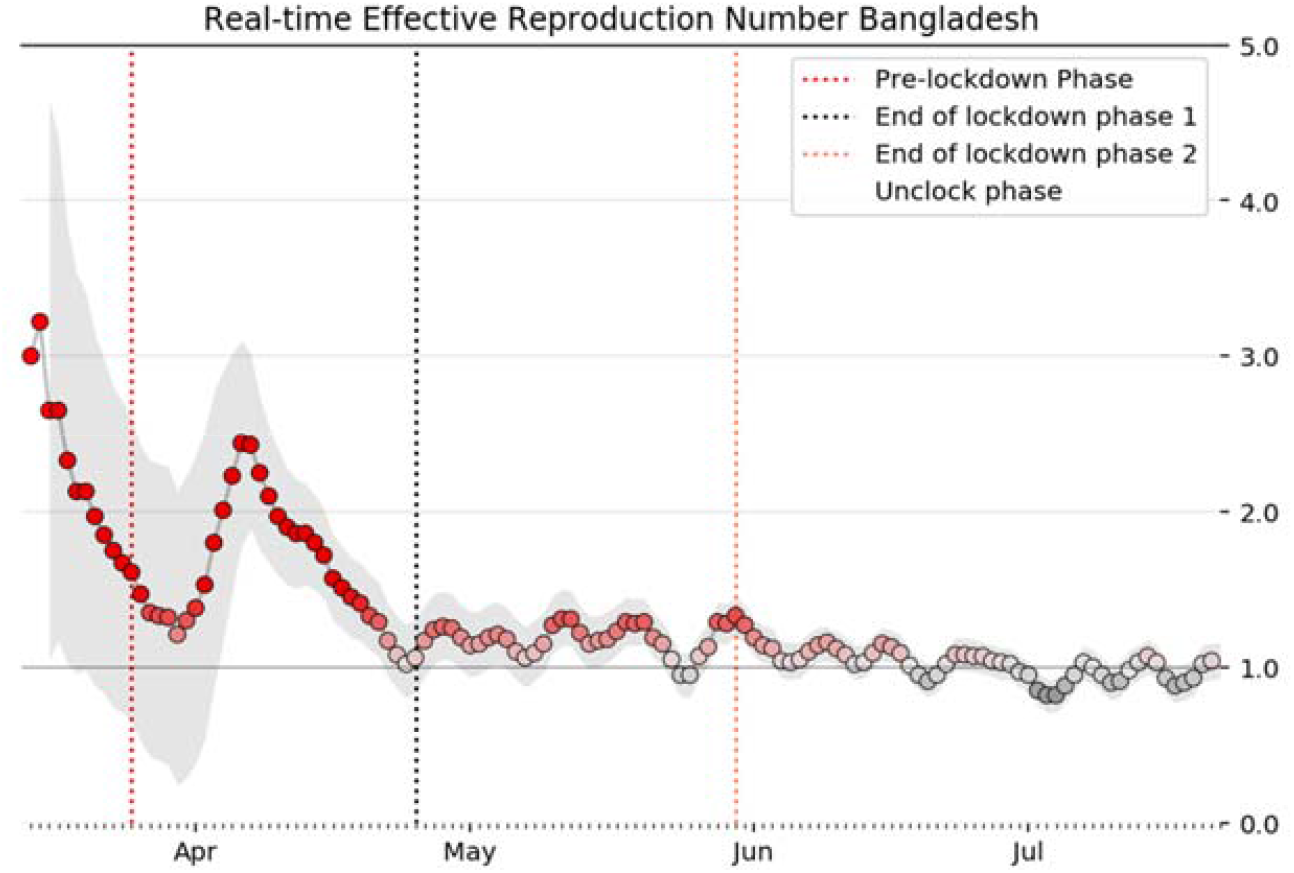
Real-time effective reproduction number (Rt) Bangladesh.

Table 2 suggests, each phase of lockdown reduced the mean Rt by 28.44%, and 26.70%, respectively. The mean Rt fell by 13.55% from May 31 to July 21, 2020, despite easing of lockdown in Bangladesh. A smoothed 7 days moving average of new cases depicts that the number of new cases started to drop at the beginning of July (Figure 2). Also, a rolling average of the testing volume was assessed to check whether testing volume had any impact on estimated Rt less than 1. It is observed that COVID-19 testing volume and the number of new cases ramped down during these periods (Figure 2, Figure 3). Thus, low testing volumes might have had an impact on underestimating lower Rt.

**Figure 2:**
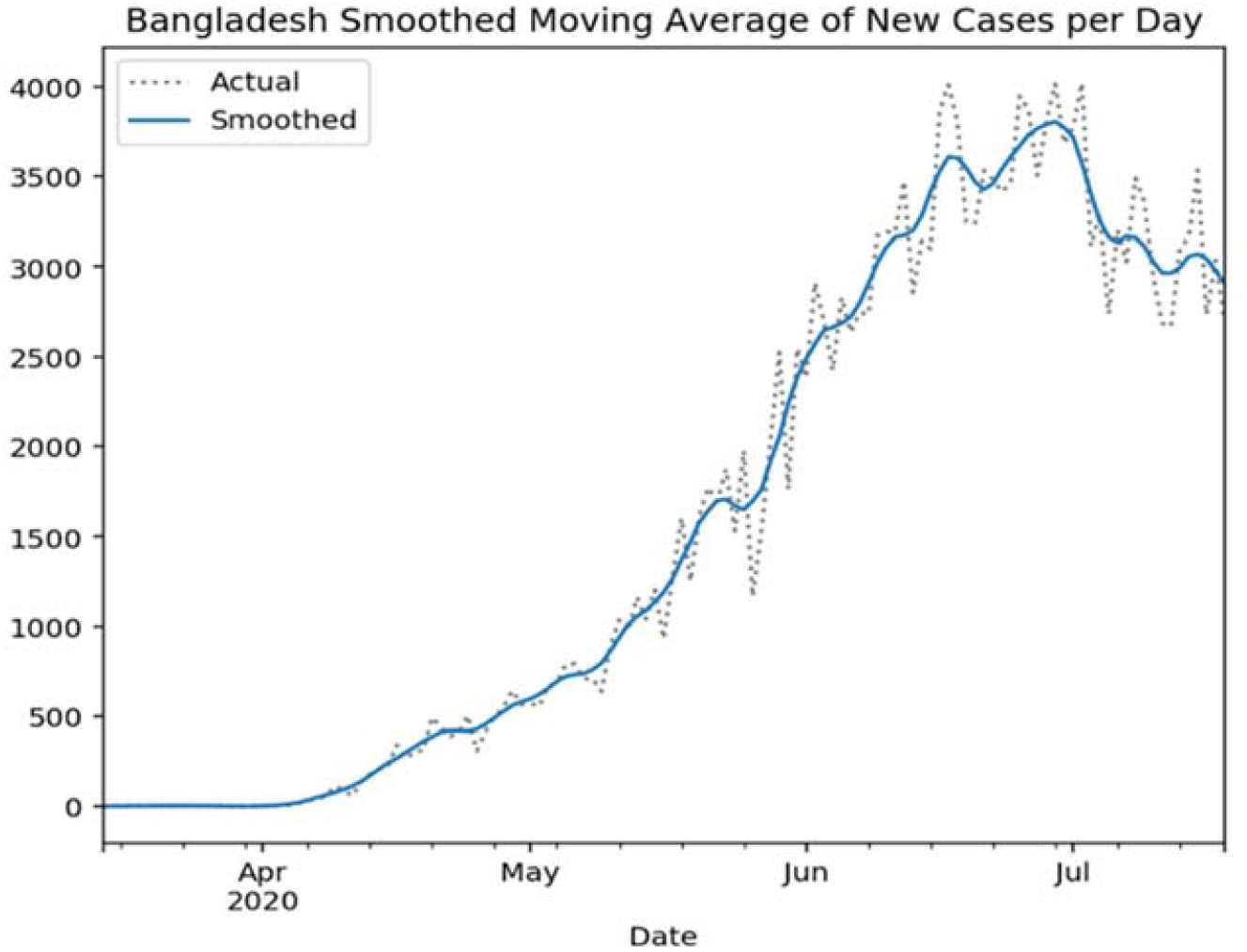
Bangladesh smoothed moving average of new cases per day.

**Figure 3:**
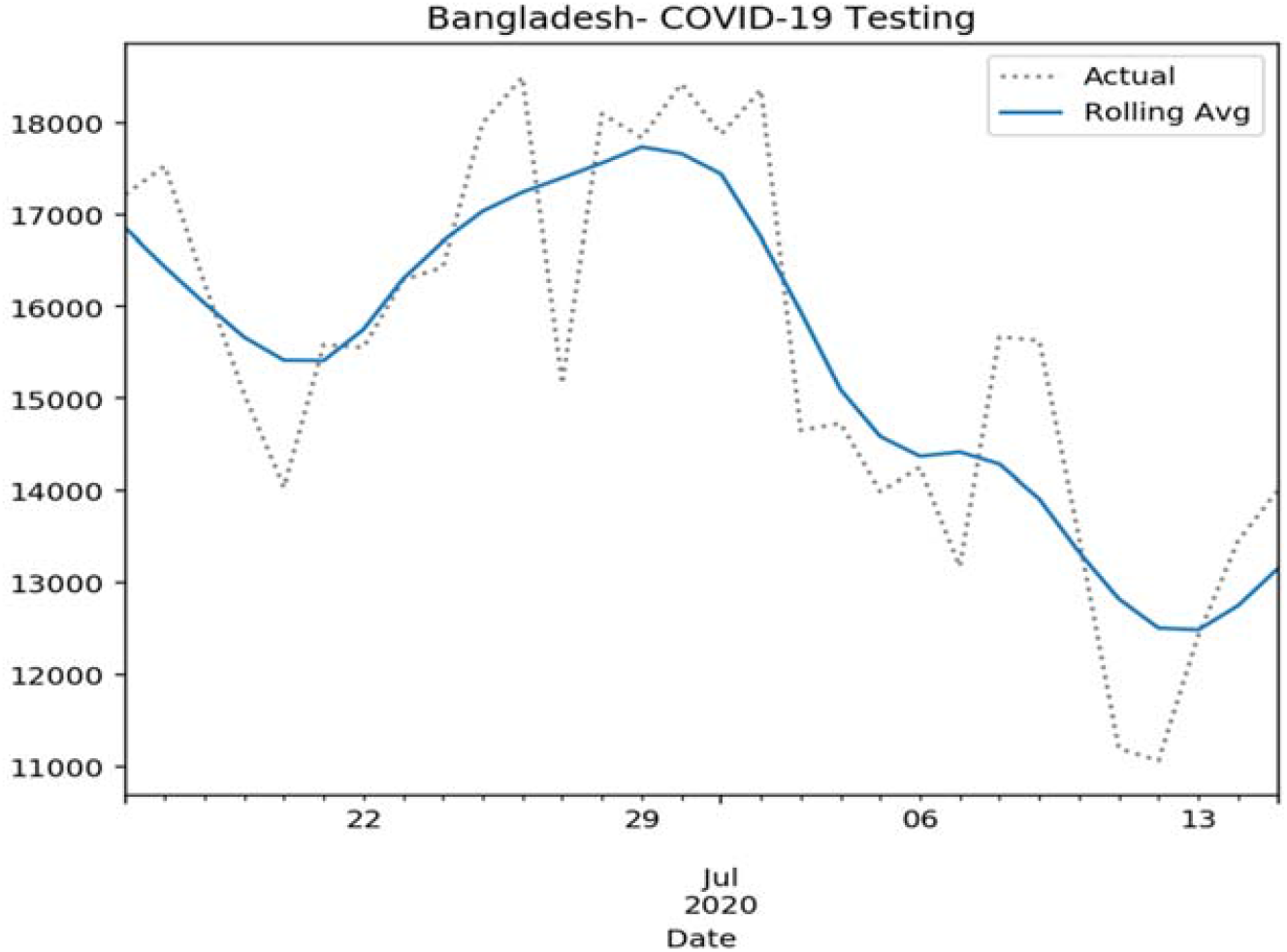
COVID-19 testing rolling average.

**Figure 4:**
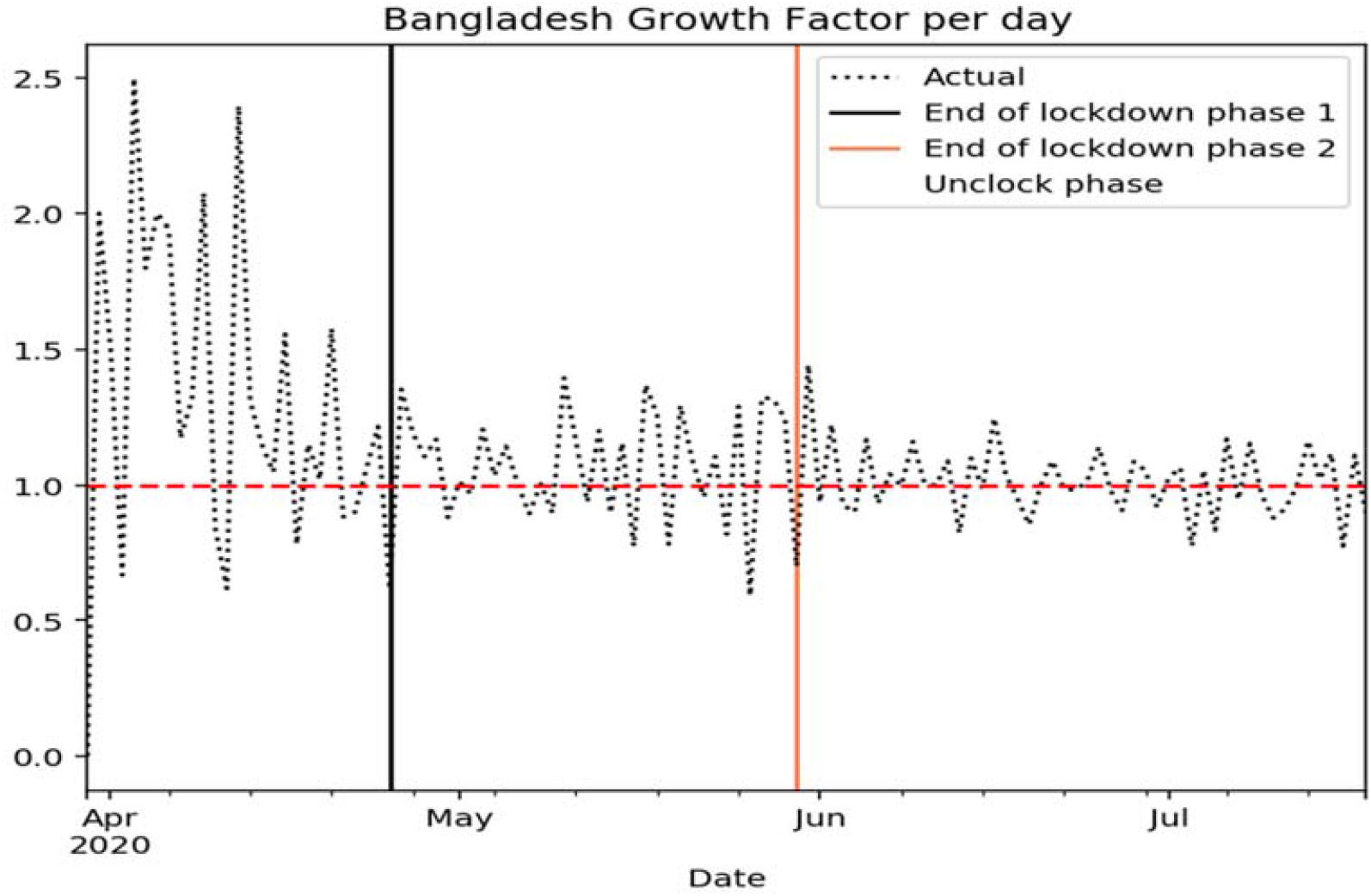
Bangladesh growth factor per day.

**Figure 5:**
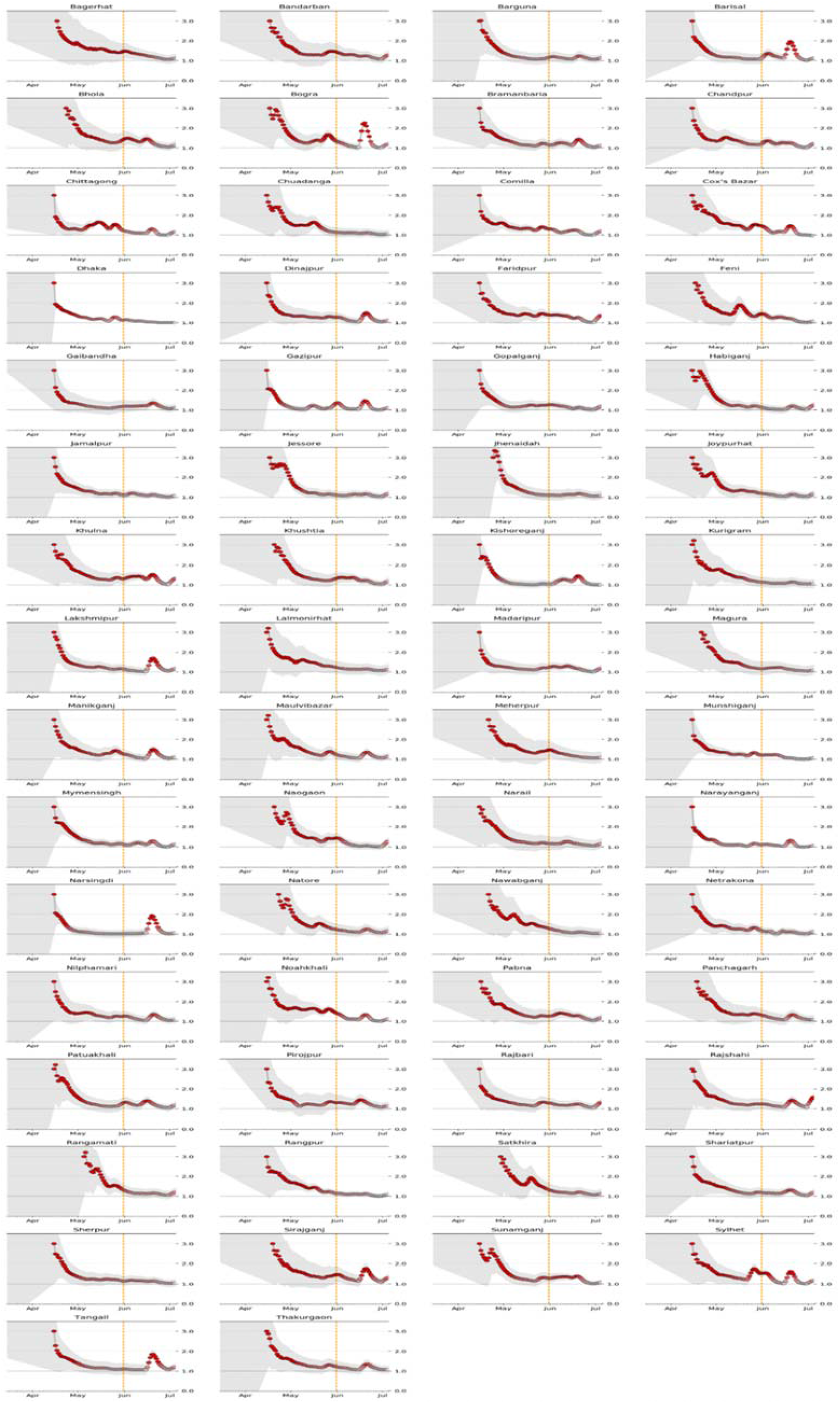
Effective reproduction number district-wise: The orange line depicts the end of the lockdown nation-wide.

### Growth Factor

It is observed that the growth factor (GF) reached a maximum of 2.5 on April 3 and during the lockdown phase 1(from 26^th^ March, 2020 to 25^th^ April, 2020) the GF was within the range of 0-2.5, while in the lockdown phase 2 (from 26^th^ April 2020 to 30^th^ May 2020), The GF fluctuated within the range of 0.70-1.39. On the 31^st^ May 2020 government of Bangladesh lifted the nation-wide lockdown. In the unlock phase (31^st^ May 2020 – 18^th^ July 2020), the GF fluctuated within the range of 0.82-1.44. The mean growth factor before imposing nation-wide lockdown was 0.66, during lockdown phase 1, GF was 1.16(a 75 % increase), during lockdown phase 2, GF was 1.07 (an 8% decrease), and during the unlock phase, GF was 1.02 (a 4.67% decrease). The mean growth factor for Bangladesh is 1.02 (from March 8, 2020, to July 21, 2020).

### District wise Rt estimation

Since the value of Rt varies spatially due to various containment strategies(e.g., maintain social distancing), in this study, we considered estimating Rt at the district-level, which will help to assess the impacts of containment strategies during lockdown period. In an epidemic like COVID-19, it is difficult to get initial stage data from sub-national or districts, because of under-reporting or delay in reporting of cases. To address this limitation, we did not consider reported cases at the district level for the first 4 weeks starting from March 15, 2020. The estimated Rt was higher at the initial stage due to few observed cases at times. It’s observed that during the nation-wide lockdown period, the Rt of most of the districts fluctuated between 2 to 1.

## Discussion and Conclusion

In this study, we estimated the effective reproduction number for COVID-19 for Bangladesh and its districts, which is a key metric for developing and assessing the containment strategies. Since the effective reproduction number varies spatially due to various factors, it supports district-wise analyses that are essential to capture the trend at the local level. Therefore, we also study the trend of Rt at the district level. In addition, under-reporting or delay in reporting as well as lack of adequate testing, may also contribute to an underestimate or overestimate of Rt estimates. Based on the results, the effective reproduction number (Rt) of Bangladesh and most of its districts is still above 1, indicating the outbreak of COVID-19 will continue to grow. The effects of lockdown at district level varied place to place, and it is observed that most of the districts’ Rt had increased after lifting nation-wide lockdown and remained above Rt > 1 till now.

It is observed that the average Rt for districts was within the range between 1.20 (1.37-1.07: 90% HDI) and 1.53 (2.27-0.92: 90% HDI), which is presented in Figure 6. The rightmost districts had higher average Rt values.

**Figure 6:**
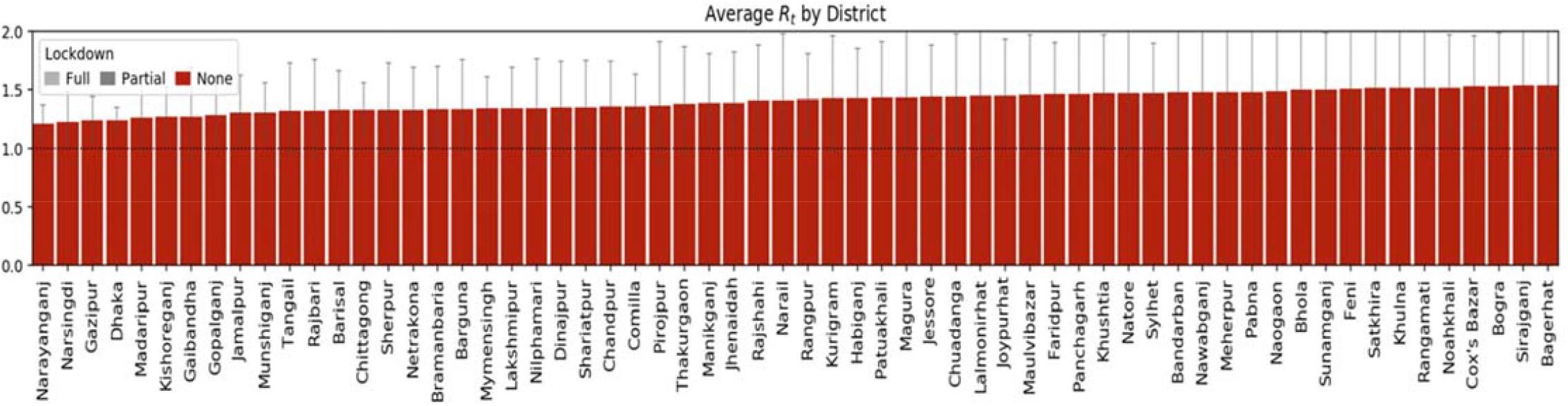
Average effective reproduction number by district.

To assess which districts had least under control, average lower limits of HDI (Low_90 HDI) of districts were assessed, which indicates the true value of Rt was certainly almost above threshold value 1. Thirty-four districts had the highest lower HDI (Figure 7), indicating these districts were not under control.According to the study, most districts indicated a significant increase of RT during June 15-30, 2020 and as of now (July 4^th^, 2020), no districts managed to take Rt below 1, which indicates cases may increase over time at the district level.

**Figure 7:**
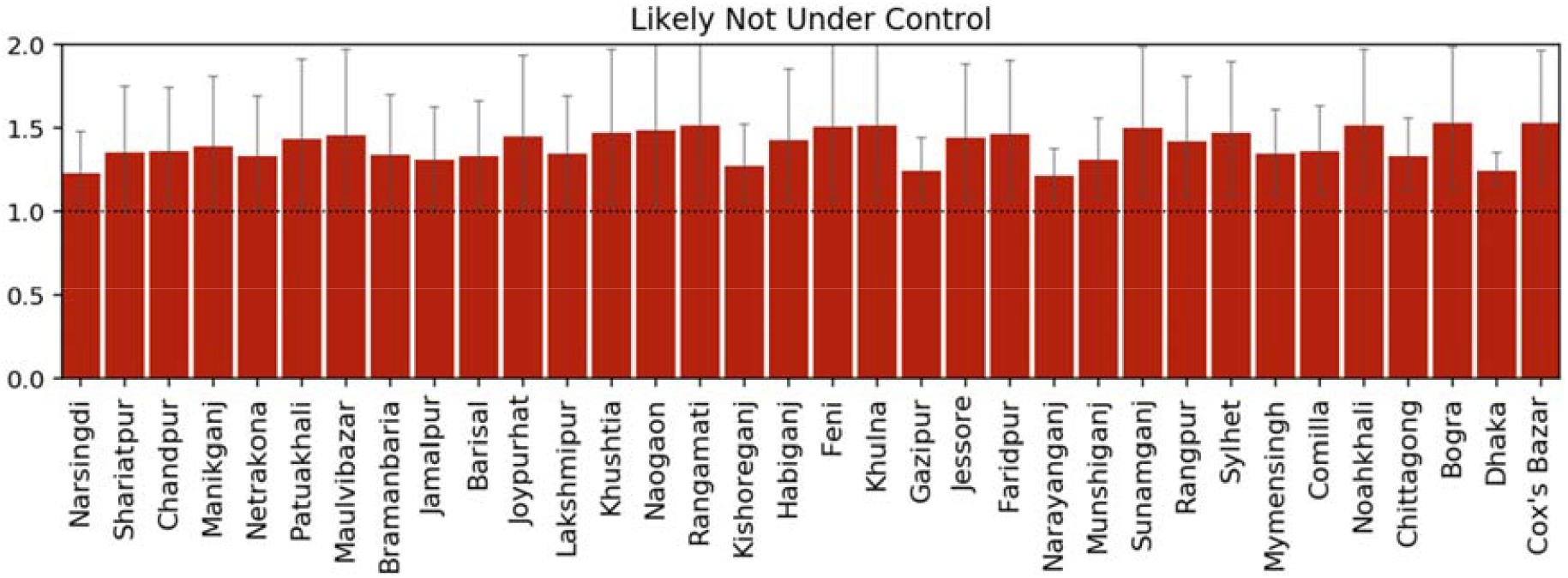
Average maximum lower HDI for districts.

The most recent Rt, before easing nation-wide lockdown Figure 8 depicts Rt ranged 1.00-1.10 for 4 districts (Narsingdi, Kishoreganj, Barisal, Jhenaidah), Rt ranged 1.11-1.20 for 21 districts (Tangail, Jamalpur, Jessore, Narayanganj, Chuadanga, Barguna, Dhaka, Kurigram, Habiganj, Rangpur, Sherpur, Mymensingh, Magura, Lakshmipur, Chandpur, Gaibandha, Bramanbaria, Narail, Netrakona, Joypurhat, Shariatpur), Rt ranged 1.21-1.30 for 19 districts (Munshiganj, Madaripur, Rajshahi, Nilphamari, Thakurgaon, Gopalganj, Chittagong, Natore, Patuakhali, Pabna, Lalmonirhat, Maulvibazar, Dinajpur, Khushtia, Manikganj, Sunamganj, Gazipur, Rajbari, Khulna), Rt ranged 1.31-1.52 for 18 districts (Pirojpur, Nawabganj, Comilla, Panchagarh, Rangamati, Satkhira, Bhola, Bogra, Faridpur, Naogaon, Sirajganj, Bagerhat, Feni, Meherpur, Bandarban, Noahkhali, Cox’s Bazar, Sylhet).

**Figure 8:**
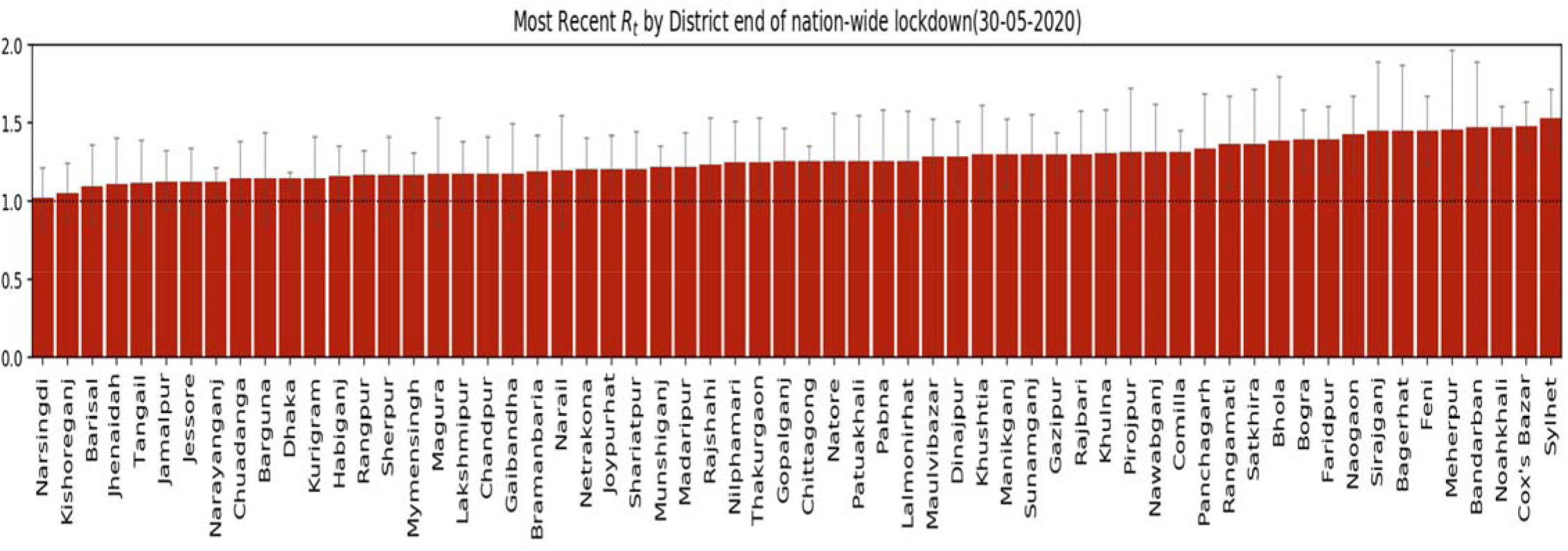
Most recent Rt by district end of the nation-wide lockdown.

During the unlock-phase (31^st^ May 2020 – 4^th^ July 2020), the most recent Rt (as of 4^th^ July 2020) by district-wise depicted on Figure 9, which suggests ten districts had the highest Rt namely, Rangamati, Chandpur, Habiganj, Naogaon, Bandarban, Sirajganj, Rajbari, Khulna, Faridpur and Rajshahi, where Rt ranged from 1.20 (Rangamati) to 1.57 (Rajshahi). The Rt was stable or Rt=1 for Cox’s Bazar, Dhaka, and Kishoreganj.

**Figure 9:**
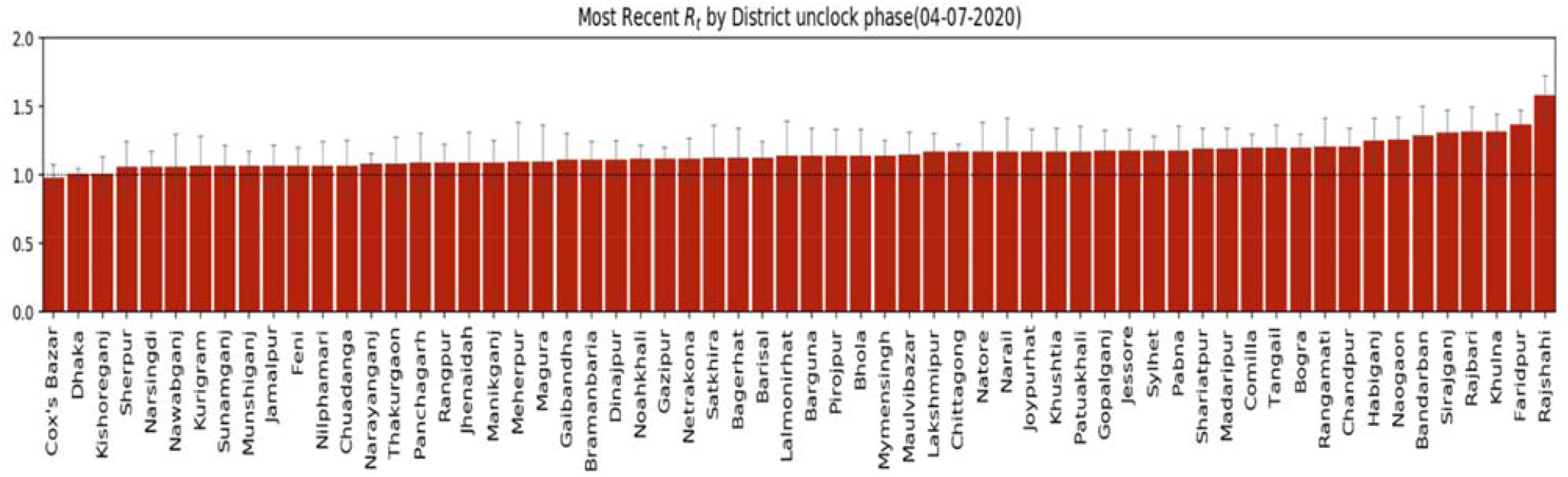
Most recent Rt by the district as of 4th July 2020.

Considering the serial time interval, the data are limited to online reports of confirmed cases and could, therefore, be biased towards more severe cases in areas with high-functioning healthcare and public health infrastructure. The rapid isolation of such cases may have prevented longer serial intervals, potentially shifting our estimate downwards compared to serial intervals that might be observed in an uncontrolled epidemic.^15^ It is estimated that if the effective reproduction number could be brought down to less than 1.5, a higher level of control can be achieved where 50% of contacts have to be traced successfully, while for effective reproduction number 2.5 and 3.5, more than 70% and 90%, of contacts, respectively, had to be traced. ^25^

To summarize, the COVID-19 outbreak represents a real threat to low-income countries like Bangladesh. The Rt estimation of the COVID-19 outbreak in Bangladesh and its district suggests an abyss scenario. This study estimated effective reproduction number Rt and growth factor since the onset of the outbreak. This study estimated mean Rt for Bangladesh is 1.32, meaning that infected people have spread the virus to an average of another person, with 0.32 added chance of infecting a second individual. The Rt of Bangladesh had dropped below 1 during May 24-25, June 19-21, from June 30 to July 6, and July 16-19,2020. We find that number of testing conducted per day might have had an impact on Rt. Thus, broader testing is necessary to assess a more realistic estimation of Rt, which will reveal the upcoming COVID-19 scenario in Bangladesh and may work as a guiding light for policy- makers. Each phase of nation-wide lockdown has contributed to the decline of effective reproduction number (Rt) for Bangladesh and reduced the mean Rt by 28.44% and 26.70%, respectively, but at districts level effects of lockdown remained uncertain. Rt started to decline monotonically from 13th week to 17th week. At districts level, 34 districts were identified as not were under control. The mean Rt for districts was within the range between 1.20 and 1.53(as of July 4, 2020). As of July 4, 2020, only districts (Cox’s Bazar, Dhaka, Kishoreganj) had stable Rt =1, and most of the districts Rt value are still above one, which indicates an exponential increase of cases may occur at these districts. The Rt on the reopening day (31^st^ May 2020) had a frightening number of 1.27, as of July 21, 2020, the most recent Rt of Bangladesh is 1.07(0.92-1.15: 90% HDI), meaning an infected person infecting 1.07 others on average, which is the slow speed of infection but still suggests the number of cases will continue to grow. Thus, it is recommended that Bangladesh should increase the number of testing per day and take necessary measures to keep Rt long way below the crucial threshold 1. Since there is no proven vaccine developed for COVID-19 yet and absent an indefinite lockdown in Bangladesh due to its high socioeconomic cost, it will be very difficult for Bangladesh to get Rt a long way below 1. Moreover, there will always be new imported cases, local outbreaks. Hence, a careful calibration of measures is needed, where Rt will shed some light on the COVID-19 situation in Bangladesh.

### Limitations

Comparatively to other Rt estimation models, adapted Bettencourt and Ribeiro methods ^22,23^ can lead to biased Rt estimates when the underlying dynamics of a pathogen are not well understood as reported by Gostic et al.^26^ The model may face limitations associated with data uncertainties such as differences in testing, inconsistent, delayed reporting, and low number cases, and high variability at the district level. It is worth noting that there is a delayed, inconsistent, and lower infection report of cases at the district level, that might overestimate Rt at the district level. The results are impacted by testing rate, increases, and decreases in the testing rate will increase and decrease reproduction number estimates, respectively. However, calculating an approximate Rt using available data may provide valuable insights into the dynamics of the COVID-19 outbreak in Bangladesh.

### Follow up

As the COVID-19 pandemic is already continuing in Bangladesh, despite no nation-wide lockdown in place, schools, college, universities remained close till now, and it is important to watch daily Rt for Bangladesh to set up comprehensive strategies by policy-makers. Given the urgency of the COVID-19 situation in Bangladesh, a daily Rt estimation for COVID-19 in Bangladesh can be found at (https://wolf-dev.github.io/rt/).

## Data Availability

All data were collected from public database and are with the corresponding author. Data will be available on a valid request.

## Acknowledgement

The authors would like to thanks Mr. Ishayat Ahmed for collecting data for this research.

## Authors Contribution

All authors contributed equally to this research.

## Conflict of interest

Authors declare that there is no conflict of interest.

## Funding

Authors did not receive any fund for this research.

## Notes

### Competing Interest Statement

The authors have declared no competing interest.

